# Offline Contextual Bandits for Lung Donor–Recipient Matching: A Retrospective Feasibility Study

**DOI:** 10.64898/2026.09.22.26363700

**Authors:** Yifan Xiang, Baraa Abed, Malvern Madondo, Navin Vigeshwar, Rishikesan Kamaleswaran, Kunal Patel

**Affiliations:** Department of Biomedical Engineering, Duke University Pratt School of Engineering, Durham, NC, USA; Department of Electrical and Computer Engineering, Duke University Pratt School of Engineering, Durham, NC, USA; Department of Surgery, Duke University School of Medicine, Durham, NC, USA; Department of Anesthesiology, Duke University School of Medicine, Durham, NC, USA

**Keywords:** Lung transplantation, Donor–recipient matching, Organ allocation, Offline Contextual bandits, Off-policy evaluation, Composite Allocation Score

## Abstract

**Background:** Lung donor–recipient matching requires balancing recipient medical urgency with expected post-transplant benefit. Conventional prediction models estimate outcomes for historical donor–recipient pairs but do not directly compare alternative candidates for the same donor. We developed an offline contextual-bandit framework to assess the feasibility of learning donor-centered lung-matching policies from historical registry data.

**Methods:** We used linked United Network for Organ Sharing (UNOS)/Organ Procurement and Transplantation Network (OPTN) Standard Transplant Analysis and Research files to identify 8,529 lung transplant allocation events from 2023 through 2025. Each event included the historical recipient and nine date-matched, ABO-compatible pseudo-candidates. The reward combined 72-hour respiratory-support-free survival (weight, 0.75) with a normalized initial Composite Allocation Score waitlist medical-urgency score (weight, 0.25). We trained a pessimistic neural lower-confidence-bound contextual-bandit policy (NeuraLCB) and evaluated it on held-out donor events using matched-action evaluation and self-normalized inverse propensity scoring (SNIPS).

**Results:** The reconstructed dataset contained 85,290 donor–candidate rows. In the held-out set of 1,706 allocation events, the observed clinician reward was 0.534 (95% CI, 0.518–0.549). NeuraLCB agreed with the historical recipient in 174 events (10.2%), with a matched-action reward of 0.616. The SNIPS-estimated value of the NeuraLCB policy was 0.649 (95% CI, 0.568–0.717), corresponding to a paired difference of +0.116 (95% CI, +0.035 to +0.181) relative to observed clinician reward. However, the effective sample size was 51.5 events, indicating limited overlap between the learned and estimated historical policies.

**Conclusions:** This study demonstrates the feasibility of constructing and evaluating an offline contextual-bandit framework for lung donor–recipient matching using registry data. The estimated policy value was higher than the observed historical reward, but this finding is exploratory because candidate sets were reconstructed and off-policy evaluation had limited overlap. Verified offer sets, time-stamped candidate data, explicit compatibility constraints, and prospective evaluation are required before clinical use.

## 1 Introduction

Lung transplantation is the definitive treatment for selected patients with end-stage lung disease, but suitable donor lungs remain scarce [1]. Allocation decisions require simultaneous consideration of donor physiology and organ quality, recipient urgency and expected benefit, blood-group and size compatibility, immunologic risk, and the practical constraints of organ offers [2]. These interacting considerations are difficult to fully encode in a single rule-based score or in a conventional outcome-prediction model.

Machine-learning models have been used to estimate post-transplant outcomes from donor, recipient, and transplant characteristics [3, 4, 5]. Such models are useful for risk prediction, but a prediction model does not itself resolve the allocation problem: for one donor, a decision-maker must compare several potential recipients and select one. Moreover, historical transplant registries observe the outcome only for the recipient who actually received the organ; the outcome that would have occurred for other potential recipients is counterfactual and unobserved [6, 7].

Offline contextual bandits provide a framework for learning from such logged decisions without intervening in clinical care [6, 8]. The donor represents the context, candidate recipients represent the available actions, the historical transplant is the logged action, and the observed post-transplant outcome defines the reward. A clinically appropriate offline policy should also account for uncertainty, because it should not make confident recommendations for donor–recipient combinations that are poorly represented in the historical data [8, 9].

In this retrospective feasibility study, we develop a donor-centered offline contextual-bandit pipeline for lung donor– recipient matching. We reconstruct candidate sets from historical data, train a pessimistic neural policy that trades off predicted reward against uncertainty, and assess the learned policy using held-out off-policy evaluation. The goal is methodological feasibility, not a clinical allocation recommendation. In particular, the reconstructed sets are not verified historical offer sets, and all findings should be interpreted as exploratory.

This work is also preliminary to our broader research program on safety-constrained donor-recipient compatibility. The program seeks to determine whether donor biological reserve and donor-recipient compatibility, rather than chronological age alone, can support more precise use of extended-criteria donor lungs. The present analysis establishes the clinical-data and offline-learning foundation for that objective: it tests whether a donor-centered ranking policy can be learned from partial historical feedback using only pre-transplant information. Future extensions will incorporate verified offer sets and hard clinical eligibility constraints, including ABO compatibility, size matching, and unacceptable-antigen restrictions, and will evaluate whether molecular measures of donor reserve add value beyond standard donor and recipient characteristics.

## 2 Methods

### 2.1 Problem Formulation

We formulate this problem as an offline contextual bandit problem, in which we have a dataset 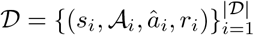, where *s*_*i*_ *∈ S* denotes the context observed for allocation event *i*, 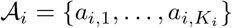denotes the set of candidate actions available for that event, *â* _*i*_ *∈ A* _*i*_ denotes the action selected by the historical behavior policy, and *r*_*i*_ *∈* ℝ denotes the observed reward resulting from the selected action. Importantly, the reward is observed only for *â*_*i*_; the counterfactual rewards associated with the remaining actions in *A*_*i*_ are unobserved.

In our setting, the context *s*_*i*_ contains features describing the donor, while each action *a*_*i,j*_ is represented by features describing a candidate recipient. We therefore represent each context–action pair by the concatenated feature vector *x*_*i,j*_ = [*s*_*i*_; *a*_*i,j*_] *∈* ℝ ^*d*^, and model the expected reward associated with a context–action pair as

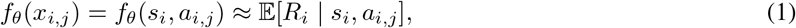

where *f*_*θ*_ is a neural reward model parameterized by *θ*.

The objective is to learn a policy *π* from the offline data that maps each context and its candidate action set to an action, *π*(*s*_*i*_, *A* _*i*_) *∈ A* _*i*_, such that the expected reward under the learned policy is maximized. Because the dataset contains outcomes only for actions selected by the historical policy, learning must account for uncertainty in context–action regions that are weakly represented in the logged data. We therefore adopt a pessimistic offline contextual-bandit formulation, in which candidate actions are evaluated using both their estimated reward and the uncertainty associated with that estimate [9].

### 2.2 Dataset Construction

#### 2.2.1 Study cohort and allocation events

We conducted a retrospective cohort study using data from the United Network for Organ Sharing (UNOS)/Organ Procurement and Transplantation Network (OPTN) Standard Transplant Analysis and Research (STAR) files[10]. We linked thoracic transplant, deceased-donor, and recipient waitlist records using the available donor and recipient identifiers. The analytic data included donor demographic and clinical characteristics; recipient demographic, waitlist,and pre-transplant clinical characteristics; allocation-related variables; transplant information; and early post-transplant respiratory-support outcomes. We restricted the cohort to lung transplant events occurring from 2023 through 2025, when the Composite Allocation Score (CAS) was available.

Each historical donor-recipient transplant with a valid donor identifier, recipient identifier, and observed 72-hour respiratory-support outcome was treated as one donor allocation event. The final analytic cohort contained 8,529 allocation events. For each event, the historically transplanted recipient was defined as the logged action, and the observed post-transplant outcome for that recipient provided the observed reward.

#### 2.2.2 Candidate-set reconstruction

For each donor event, we created a set of 10 candidate recipients. The set always included the recipient who historically received the donor lung, designated the logged action. We selected nine pseudo-candidates from recipients transplanted in the same dataset split, within a prespecified calendar-time window around the donor event, and satisfying the ABO-compatibility rule. Pseudo-candidates were sampled without replacement within an event. This procedure produced 85,290 donor–candidate rows: 8,529 logged recipient rows and 76,761 pseudo-candidate rows.

Note that these sets are reconstructed decision sets, not verified match-run offer sets. Therefore, a pseudo-candidate may not have been active on the waitlist, eligible, offered the organ, or willing to accept it at the historical allocation time.

#### 2.2.3 Feature Selection and Preprocessing

Feature selection was guided by clinical relevance, temporal availability, and whether a variable could be defined consistently for every donor-candidate pair in a reconstructed candidate slate. We retained donor variables describing organ quality and physiology, including demographics, anthropometrics, blood group, oxygenation and arterial blood-gas measures, imaging and bronchoscopy findings, smoking and substance-use history, comorbidities, infectious-risk indicators, cause of death, cardiopulmonary arrest and resuscitation history, hemodynamics, and vasoactive or ventilatory support. Recipient variables described demographics, body size, blood group, pulmonary diagnosis, medical urgency, functional status, respiratory and extracorporeal support, oxygen requirement, renal function and dialysis, pulmonary function, hemodynamics, prior transplantation or thoracic surgery, and sensitization measures when available.

We excluded identifiers, audit fields, administrative variables, and variables with no observed values. Variables were also excluded when they could not plausibly have been available at the time of allocation, including post-transplant treatments and outcomes. Variables with greater than 80% missingness were removed.

Numerical features were median-imputed, augmented with missingness indicators, and standardized using the training data. Categorical features were assigned an explicit missing category and one-hot encoded. All preprocessing parameters, including medians, scaling factors, categorical levels, and the set of retained variables, were estimated using the training data and then applied unchanged to subsequent data. This procedure prevents leakage of the evaluation set.

The same donor-level representation was assigned to each candidate within a donor event, whereas recipient-level features varied across candidates. Candidate selection was limited to the historically transplanted recipient and date-matched, ABO-compatible pseudo-candidates. Because verified offer-level information was unavailable, the reconstructed sets did not incorporate center-specific acceptance behavior, geographic distance, predicted lung-size compatibility, HLA or unacceptable-antigen compatibility, or exact crossmatch eligibility. These factors should be incorporated in future analyses using verified match-run offer sets and time-stamped candidate data.

#### 2.2.4 Reward definition

The reward was defined only for the logged recipient and combined short-term post-transplant respiratory recovery with pre-transplant medical urgency. Let *Y*_*i*_ = 1 when the recipient survived without extracorporeal membrane oxygenation and without invasive mechanical ventilation 72 hours after transplantation, and *Y*_*i*_ = 0 otherwise. Let *C*_*i*_ denote the recipient’s initial CAS waitlist medical-urgency score, rescaled to [0, 1] by min–max normalization among eligible logged recipients. The composite reward was

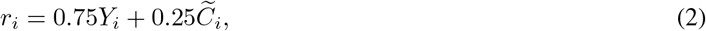

where 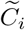is the normalized urgency score. Thus, respiratory-support-free survival is the dominant component, while the CAS term gives additional weight to allocation to recipients with greater measured medical urgency. This composite reward ranges from 0 to 1 and balances recipient urgency and post-transplant benefit.

### 2.3 Experimental Details

#### 2.3.1 Algorithm

We adapt NeuraLCB [9], a pessimistic neural contextual bandit algorithm, to the donor–recipient allocation setting. The algorithm consists of a neural reward model and an uncertainty estimator defined in the neural parameter space. For each donor context *s*_*i*_ and candidate recipient *a*_*i,j*_, we construct the context–action representation *x*_*i,j*_ = [*s*_*i*_; *a*_*i,j*_], and use a neural network *f*_*θ*_ to estimate the expected reward, 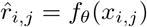.

To quantify uncertainty in the reward prediction, NeuraLCB uses the gradient of the neural network with respect to its parameters. At iteration *t*, we define

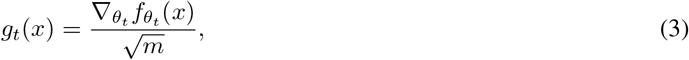

where *θ*_*t*_ denotes the current neural-network parameters and *m* denotes the network width. Following the approximate variant of NeuraLCB, we maintain a diagonal approximation to the confidence matrix. In the original implementation, a separate confidence matrix is maintained for each discrete action. In our setting, actions correspond to recipients represented by their own feature vectors rather than a fixed set of action identities. We therefore maintain a single shared diagonal confidence matrix,

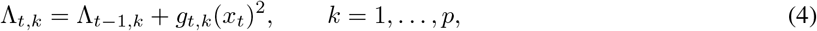

where *x*_*t*_ = [*s*_*t*_; *â*_*t*_] denotes the historically observed donor–recipient pair at iteration *t*. The confidence state is initialized as Λ_0_ = *λ*_0_**1**. This allows uncertainty to be represented in the shared neural parameter space across donor–recipient pairs.

For a candidate pair *x*, the resulting uncertainty estimate is

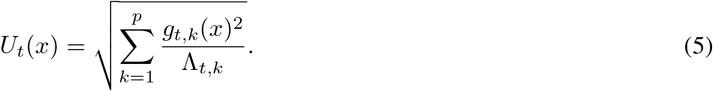

Candidate recipients are scored pessimistically according to the lower confidence bound

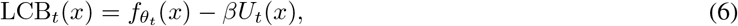

and the recipient with the largest LCB score is selected,

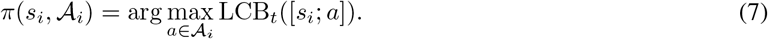

Thus, the policy favors candidates with high predicted reward while penalizing predictions associated with greater uncertainty under the logged data.

#### 2.3.2 Evaluation

We evaluate the learned policy using held-out allocation events that are not used during training. Because rewards are observed only for the recipient selected by the historical clinician policy, we first perform matched-action (replay) evaluation. For each test event, NeuraLCB scores all candidate recipients and selects the candidate with the largest LCB. When this selection agrees with the historically selected recipient, the corresponding observed reward is available for evaluation. We report the mean observed reward among these matched events together with policy coverage, defined as the proportion of test events for which the learned and historical policies agree.

Because matched-action evaluation discards events on which the policies disagree, we additionally estimate policy value using self-normalized inverse propensity scoring (SNIPS). The historical clinician propensities are not observed in the dataset and are therefore estimated using a behavior-cloning model. The behavior model applies a shared neural scorer to each donor–recipient pair within an allocation event and normalizes the resulting logits across candidate recipients using a softmax,

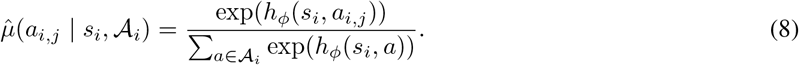

The behavior model is trained using multiclass cross-entropy to predict the recipient historically selected by the clinician.

For a target policy *π*, the importance weight associated with logged event *i* is

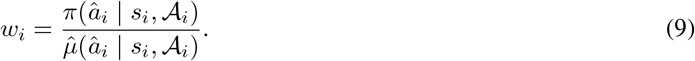

Because NeuraLCB defines a deterministic target policy, it assigns probability 1 to the action selected by NeuraLCB and 0 to all other candidate actions. We then estimate policy value using

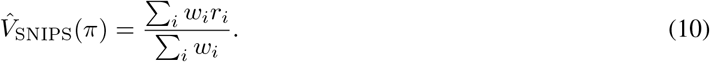

We report the effective sample size of the importance weights as an overlap diagnostic and obtain 95% confidence intervals using nonparametric bootstrap resampling at the allocation-event level. Differences between the estimated NeuraLCB policy value and the observed historical-policy reward are evaluated using paired bootstrap confidence intervals.

Finally, we analyze the learned reward function to characterize the relationships captured by the model. We use SHAP values to quantify the contribution of individual donor and recipient characteristics to predicted reward. We additionally perform controlled sensitivity analyses for donor–recipient age and BMI differences. In these analyses, all model inputs are held fixed except recipient age or BMI, which is systematically varied relative to the corresponding donor characteristic. For each specified difference, only observations for which the intervention lies within the prespecified valid range are included, and the predicted reward is compared with the corresponding donor–recipient matched reference for the same observations using paired comparisons. These analyses characterize the sensitivity of the learned reward function and are not interpreted as estimates of causal effects.

#### 2.3.3 Training and Hyperparameters

We use an 80/20 train–test split at the event level, with a fixed random seed of 42. The reward model is a multilayer perceptron with two hidden layers of width *m* = 100, each using ReLU activation. The model is trained by minimizing mean squared error with an *l*_2_ regularization coefficient of *λ* = 10^*™*4^. Optimization is performed using Adam [11] with a learning rate of 10^*™*3^. At each NeuraLCB update, the model is trained for 100 optimization steps using minibatches of size 32. The confidence parameters are set to *λ*_0_ = 0.1 and *β* = 0.1.

The behavior-cloning model consists of a single hidden layer with 64 units and ReLU activation. It is trained using Adam with a learning rate of 10^*™*3^ and multiclass cross-entropy loss. Within the training set, 20% of events are reserved for validation. We use early stopping with a patience of 25 optimization steps and a minimum validation-loss improvement of 10^*™*5^.

## 3 Results

### 3.1 Policy Evaluation

We present the off-policy evaluation results in Table 2. NeuraLCB achieved a matched-action reward of 0.616, compared with an overall observed clinician reward of 0.534. However, the learned policy agreed with the historical clinician decision in only 10.2% of test events (174 of 1,706).

**Table 1:** Characteristics of historical lung donor–recipient allocation events, stratified by 72-hour respiratory-support status.

| Model-input characteristic | Respiratory-support-free<br>at 72 h ($n = 5,720$) | ECMO and/or invasive<br>ventilation at 72 h ($n = 2,809$) | $P$ value |
| --- | --- | --- | --- |
| *Donor input features* |  |  |  |
| Age, years | 37.0 (27.0–48.0) | 39.0 (28.0–50.0) | < 0.001 |
| Female sex | 2,069 (36.2) | 1,219 (43.4) | < 0.001 |
| Height, cm | 172.7 (165.0–178.0) | 170.0 (163.0–177.8) | < 0.001 |
| Weight, kg | 78.6 (67.6–91.6) | 75.8 (64.3–89.0) | < 0.001 |
| Body mass index, kg/m<sup>2</sup> | 26.4 (23.3–30.7) | 26.2 (22.8–30.4) | 0.012 |
| Blood group O | 3,055 (53.4) | 1,600 (57.0) | 0.002 |
| PaO<sub>2</sub>/FiO<sub>2</sub> ratio | 447.0 (385.3–505.0) | 445.0 (381.0–503.0) | 0.282 |
| History of cigarette use | 525 (9.2) | 242 (8.6) | 0.416 |
| History of hypertension | 1,762 (30.8) | 912 (32.5) | 0.126 |
| Legally brain dead | 4,829 (86.9) | 2,273 (83.3) | < 0.001 |
| *Recipient input features* |  |  |  |
| Age, years | 63.0 (57.0–68.0) | 62.0 (52.0–67.0) | < 0.001 |
| Female sex | 2,063 (36.1) | 1,257 (44.7) | < 0.001 |
| Height, cm | 170.2 (162.6–177.8) | 167.9 (160.0–175.5) | < 0.001 |
| Weight, kg | 75.3 (63.8–86.8) | 76.6 (64.0–88.4) | 0.046 |
| Body mass index, kg/m<sup>2</sup> | 26.0 (22.6–29.3) | 27.1 (23.3–30.2) | < 0.001 |
| Blood group O | 2,592 (45.3) | 1,276 (45.4) | 0.941 |
| Initial CAS waitlist medical-urgency score | 0.4 (0.2–1.2) | 0.6 (0.3–3.5) | < 0.001 |
| ECMO before transplantation | 262 (4.6) | 465 (16.6) | < 0.001 |
| Ventilatory support before transplantation | 579 (10.1) | 537 (19.1) | < 0.001 |
| Prior lung transplantation | 135 (2.4) | 135 (4.8) | < 0.001 |
| Serum creatinine, mg/dL | 0.8 (0.7–0.9) | 0.8 (0.6–1.0) | 0.906 |
| Forced vital capacity | 52.0 (41.0–66.0) | 48.0 (37.0–65.0) | < 0.001 |
| Calculated panel-reactive antibody, % | 0.0 (0.0–11.0) | 0.0 (0.0–19.2) | < 0.001 |

Values are median (interquartile range) or $n$ (%). $P$ values were calculated using Mann–Whitney U tests for continuous variables and chi-square tests for categorical variables. Missing values were excluded from the corresponding comparison. CAS, Composite Allocation Score; ECMO, extracorporeal membrane oxygenation.

**Table 2:** Off-policy evaluation of NeuraLCB on the held-out test set. Confidence intervals are 95% bootstrap intervals.

| Metric | Estimate | 95% CI |
| --- | --- | --- |
| Clinician observed reward | 0.534 | [0.518, 0.549] |
| Matched-action reward | 0.616 | – |
| Policy coverage | 10.2% | – |
| SNIPS | 0.649 | [0.568, 0.717] |
| Effective sample size | 51.5 / 1706 | – |

The SNIPS estimate was higher than the observed clinician reward, with a paired difference of +0.116 (95% CI: [+0.035, +0.181]). The paired bootstrap interval excludes zero, indicating a statistically significant difference under the estimated off-policy evaluation.

These results should nevertheless be interpreted cautiously. NeuraLCB agrees with the historically selected recipient in only 10.2% of test events, and the SNIPS estimate has an effective sample size of only 51.5 out of 1,706 events.

This indicates limited overlap between the learned and estimated clinician policies and reduces the reliability of the off-policy estimate.

An additional limitation arises from the construction of the candidate sets. For each event, the historically selected recipient is observed, whereas the remaining candidates are retrospectively constructed pseudo-candidates rather than recipients known to have been simultaneously considered by the clinician. Consequently, disagreement between NeuraLCB and the historical action does not necessarily represent disagreement between the two policies over the same choice set. We therefore view the SNIPS results as suggestive rather than conclusive evidence of improved policy performance.

Given these limitations, we next analyze the learned reward function through interpretability and controlled sensitivity analyses to characterize the donor–recipient relationships captured by the model.

### 3.2 SHAP Analysis

We use SHAP [12] to examine the contribution of individual donor and recipient characteristics to the learned reward function. Figure 2 shows the 25 features with the largest mean absolute SHAP values, where positive SHAP values indicate an increase in predicted reward and negative values indicate a decrease.

**Figure 1:**
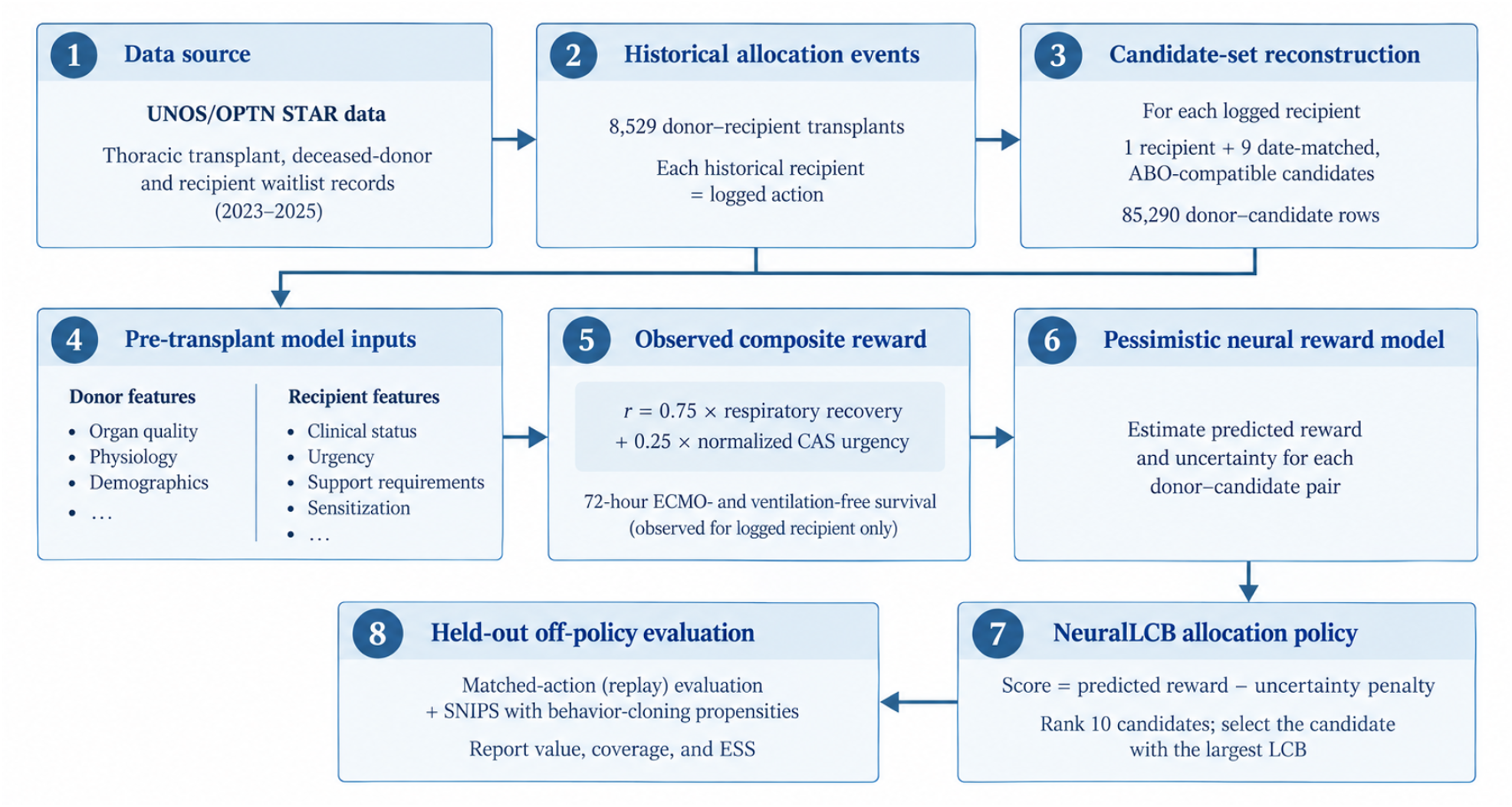
Donor-centered offline contextual-bandit framework for lung donor–recipient matching. Historical UNOS/OPTN allocation events were converted into reconstructed donor-centered candidate sets. Pre-transplant donor and recipient features were used to learn a composite reward combining 72-hour respiratory-support-free survival and normalized CAS medical urgency. A pessimistic NeuraLCB policy ranked candidates using predicted reward penalized by uncertainty, and performance was assessed using held-out off-policy evaluation.

**Figure 2:**
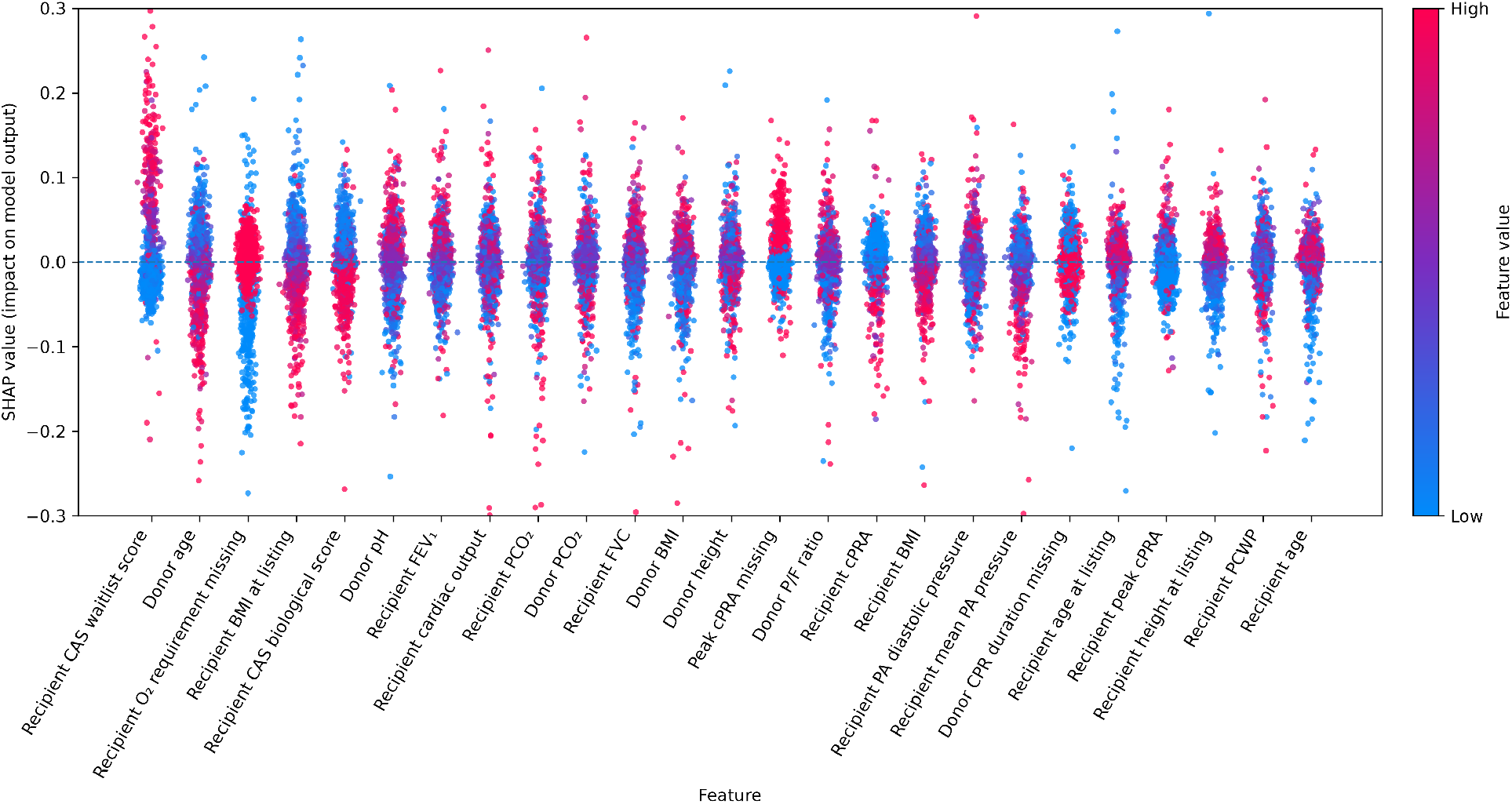
SHAP summary plot for the learned neural reward model. Features are ordered by mean absolute SHAP value. Color denotes the standardized feature value, with red indicating higher values and blue indicating lower values.

The CAS waitlist score has the largest contribution to the learned reward. This is expected because the waitlist score is explicitly included as a component of the reward, allowing the allocation objective to account for recipient urgency in addition to post-transplant outcomes. Accordingly, its prominence in the SHAP analysis primarily reflects the construction of the reward rather than a newly learned association.

Several donor characteristics also exhibit clinically interpretable relationships with the learned reward. Increasing donor age is generally associated with lower predicted reward, with younger donors tending to contribute positively to the reward prediction. Donor age has previously been investigated as a potential determinant of lung-transplant outcomes, although reported effects vary across recipient populations and donor-selection criteria [13]. Donor P/F ratio shows the opposite pattern, with higher values generally contributing positively to predicted reward. As the P/F ratio reflects pulmonary oxygenation, this relationship is consistent with the model assigning greater reward to donors with better measured lung function.

Recipient characteristics also contribute substantially to the learned reward. Higher recipient BMI generally contributes negatively to predicted reward, while lower BMI tends to contribute positively. Recipient cPRA also appears among the more influential features, with higher cPRA values generally contributing negatively to predicted reward. As higher cPRA indicates greater sensitization, the learned model therefore associates greater recipient sensitization with lower reward.

Overall, the SHAP analysis indicates that the reward model uses information from both donor quality and recipient characteristics. Importantly, these SHAP values describe associations learned by the reward model and should be interpreted together with the controlled donor–recipient sensitivity analyses presented below.

### 3.3 Sensitivity Analysis

To further examine the relationships captured by the learned reward model, we performed controlled sensitivity analyses of donor–recipient age and BMI differences. For each test observation, we varied the recipient characteristic while holding all other donor and recipient features fixed. Age differences were evaluated from *™* 30 to +30 years (in 5-year increments), and BMI differences from *™* 10 to +10 kg/m^2^ (in 2.5 kg/m^2^ increments). At each specified difference, we included observations for which the modified value remained within the prespecified valid range (age: 18–80 years; BMI: 15–50 kg/m^2^). Predicted reward was compared with a matched reference in which the recipient characteristic was set equal to the corresponding donor characteristic. Figure 3 reports the mean paired change in predicted reward, with 95% confidence intervals.

**Figure 3:**
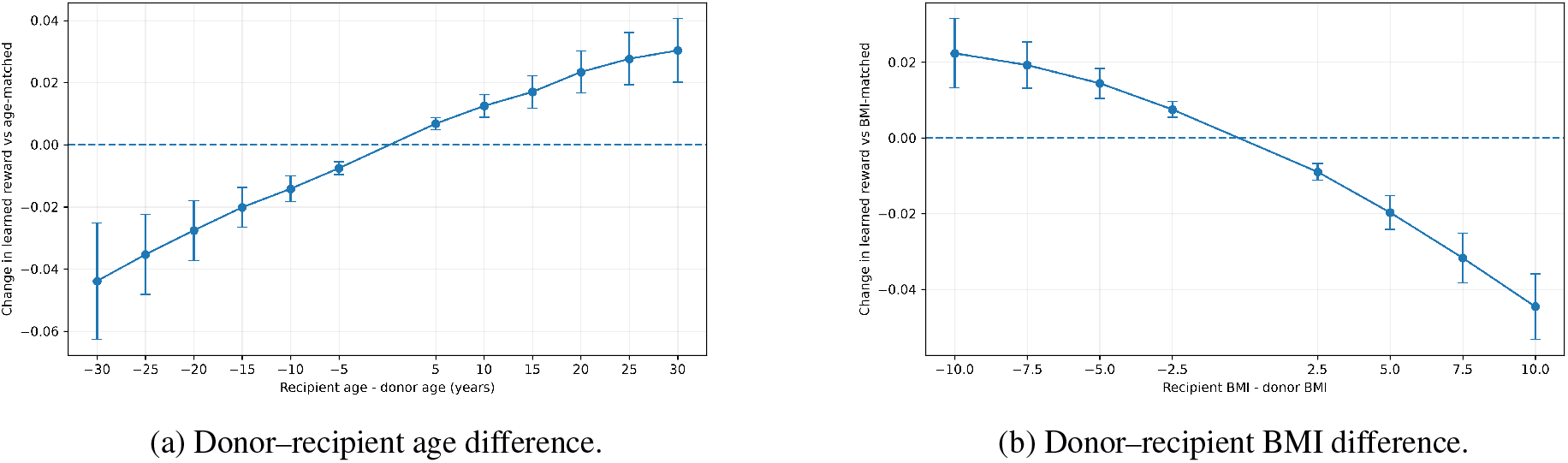
Sensitivity of the learned reward model to donor–recipient age and BMI differences. The horizontal dashed line represents the predicted reward under a matched donor–recipient reference. Error bars denote 95% confidence intervals for the mean paired change in predicted reward.

Figure 3a shows a clear asymmetric relationship between donor and recipient age. Relative to an age-matched pair, predicted reward decreases as the recipient becomes progressively younger than the donor, whereas predicted reward increases when the recipient is older than the donor. This suggests that the learned model particularly penalizes allocation of older donor lungs to younger recipients. This pattern is clinically plausible and is consistent with prior evidence suggesting that the effect of older donor age may be more pronounced among younger recipients [13].

A similarly asymmetric relationship is observed for BMI (Figure 3b). Relative to BMI-matched pairs, predicted reward increases when recipient BMI is lower than donor BMI and decreases when recipient BMI is higher than donor BMI, with the magnitude of the effect increasing as the mismatch becomes larger. This suggests that the learned reward model favors allocation from donors with higher BMI to recipients with lower BMI, while penalizing the reverse pairing. One possible interpretation is that the model has learned an asymmetric donor–recipient size relationship, whereby a relatively larger donor is more favorable than a relatively smaller donor for a given recipient.

## 4 Discussion

In this retrospective feasibility study, we developed a donor-centered offline contextual-bandit framework for lung donor–recipient matching using national transplant-registry data. The framework treats each donor as a decision context, candidate recipients as potential actions, the historical transplant as the logged action, and the observed recipient outcome as partial feedback. This formulation differs from conventional post-transplant risk prediction because it explicitly addresses the allocation question: among the candidates considered for a donor, which allocation best optimizes a prespecified clinical objective?

The reward was designed to balance post-transplant benefit and recipient urgency. Respiratory-support-free survival at 72 hours represented the dominant component of the reward, whereas the normalized initial CAS waitlist medical-urgency score gave additional weight to candidates with greater pre-transplant urgency [2]. This distinction is important: urgency is known at the time of allocation, whereas short-term post-transplant recovery is uncertain and must be learned from historical data. Including the CAS urgency component in the reward therefore specifies the clinical tradeoff that the policy is intended to optimize; it does not imply that the model independently discovers the urgency score.

The learned NeuraLCB policy had a higher SNIPS-estimated value than the observed historical clinician reward in the held-out data. The direction of this estimate is encouraging, but it should not be interpreted as evidence that the learned policy would improve clinical outcomes if deployed. First, agreement between NeuraLCB and the historical allocation was only 10.2%, indicating that the learned policy often selected a different candidate from the historical recipient. Second, the effective sample size for SNIPS was 51.5 of 1,706 held-out events. This low effective sample size indicates limited overlap between the target policy and the estimated behavior policy; consequently, the importance-weighted estimate depends on a relatively small subset of informative events and may be sensitive to behavior-model misspecification[7, 14]. The confidence interval quantifies uncertainty under the assumptions of the fitted off-policy evaluation procedure, not the certainty of a causal or clinically deployable benefit.

The candidate sets represent the most important limitation of this preliminary analysis. Each set contained the historical recipient and nine date-matched, ABO-compatible pseudo-candidates rather than the verified match-run offer set. Thus, pseudo-candidates may not have been active on the waitlist, geographically feasible, clinically eligible, offered the donor lung, or willing to accept the organ at the historical allocation time. The reconstructed sets also did not incorporate center-specific acceptance behavior, predicted lung-size compatibility, unacceptable-antigen or HLA compatibility, exact crossmatch eligibility, or anticipated ischemic time. Accordingly, disagreement between NeuraLCB and the historical action does not necessarily reflect a disagreement over the same clinically feasible choice set.

A related limitation is temporal alignment of recipient information. Although features were selected to represent donor and recipient characteristics available before transplantation, registry variables may reflect different update times across recipients. This concern is particularly relevant for pseudo-candidates, whose recorded pre-transplant variables may correspond to their own transplant event rather than the precise time of the focal donor offer. In addition, recipient urgency and several clinical variables may influence both historical allocation decisions and post-transplant outcomes, making causal interpretation difficult in observational data.

The interpretability and sensitivity analyses provide useful descriptive insight into the learned reward function but should not be interpreted causally. The prominence of the CAS waitlist score in the SHAP analysis is expected because it is incorporated directly into the composite reward [15]. The associations observed for donor age, donor oxygenation, recipient BMI, sensitization, and donor–recipient age or BMI differences describe patterns learned from the historical data. They do not establish that altering any one characteristic would change the post-transplant outcome. In particular, the controlled age and BMI analyses hold other model inputs fixed and characterize the model’s local response, not the effect of changing a donor–recipient pairing in practice.

Despite these limitations, the present work provides a foundation for the broader goal of safety-constrained donor– recipient compatibility modeling. The next stage should reconstruct verified match-run offer sets and use clinical data explicitly timestamped before each offer. Future policies should first apply hard eligibility constraints—including ABO compatibility, size compatibility, unacceptable-antigen restrictions, crossmatch feasibility, and geographic or ischemic-time constraints—before ranking clinically feasible candidates. Longer-term graft and patient outcomes, waitlist outcomes, and equity objectives should also be incorporated into a multi-objective reward framework. Ultimately, prospective silent evaluation, clinical review, and governance will be necessary before any decision-support or allocation application.

## 5 Conclusion

We developed an offline contextual-bandit framework for donor-centered lung donor–recipient matching using linked UNOS/OPTN registry data. A pessimistic neural policy produced a higher estimated composite reward than observed historical allocation in held-out evaluation, but the estimate was based on reconstructed candidate sets and showed limited policy overlap. These results establish methodological feasibility rather than clinical effectiveness. Future work using verified offer sets, time-valid recipient data, complete compatibility constraints, and prospective evaluation is required before translation to clinical decision support or organ-allocation practice.

## Supporting information

Supplemental Table S1 Model input features

## Data Availability

The data used in this study were obtained from the United Network for Organ Sharing/Organ Procurement and Transplantation Network Standard Transplant Analysis and Research files under a data-use agreement. These data are not publicly available and cannot be shared by the authors. Qualified investigators may request access directly through the Organ Procurement and Transplantation Network data-request process, subject to applicable approvals and data-use requirements. Analytic code and variable definitions will be made available from the corresponding author upon reasonable request, subject to the data-use agreement.

