## Supplemental Table S1 Model input features for "Offline Contextual Bandits for Lung Donor–Recipient Matching: A Retrospective Feasibility Study"

### Supplementary Materials

#### Model-Input Features

Table S1: Model-input features used in the offline contextual-bandit analysis.

| Domain | Feature | Input variable |
|---|---|---|
| *Donor features* | | |
| Demographic | Age, years | donor_AGE_DON |
| Demographic | Sex | donor_GENDER_DON |
| Anthropometric | Height, cm | donor_HGT_CM_DON_CALC |
| Anthropometric | Weight, kg | donor_WGT_KG_DON_CALC |
| Anthropometric | Body mass index, $kg/m^2$ | donor_BMI_DON_CALC |
| Compatibility-related | Blood group | donor_ABO_DON |
| Blood gas/oxygenation | Arterial $PaO_2$ | donor_PO2_DON |
| Blood gas/oxygenation | $PaO_2$ at recorded $FiO_2$ | donor_PO2_FIO2_DON |
| Blood gas/oxygenation | Arterial $PaCO_2$ | donor_PCO2_DON |
| Blood gas/oxygenation | Arterial pH | donor_PH_DON |
| Blood gas/oxygenation | Fraction of inspired oxygen | donor_FIO2_FRACTION |
| Blood gas/oxygenation | $PaO_2/FiO_2$ ratio | donor_PF_RATIO |
| Imaging | Chest radiograph finding | donor_CHEST_XRAY_DON |
| Bronchoscopy | Left bronchoscopy finding | donor_BRONCHO_LT_DON |
| Bronchoscopy | Right bronchoscopy finding | donor_BRONCHO_RT_DON |
| Substance-use history | Cigarette-use history | donor_HIST_CIG_DON |

*Table S1 continued*

| Domain | Feature | Input variable |
|---|---|---|
| Substance-use history | Intravenous drug-use history | `donor_HIST_IV_DRUGUSE` |
| Comorbidity | Diabetes history | `donor_HIST_DIABETES_DON` |
| Comorbidity | Hypertension history | `donor_HIST_HYPERTENS_DON` |
| Donor course | Hypertension during donor management | `donor_HYPERTENS_DUR_DON` |
| Infection | Clinical infection | `donor_CLIN_INFECT_DON` |
| Infection | Pulmonary infection | `donor_PULM_INF_DON` |
| Infection | Blood infection | `donor_BLOOD_INF_DON` |
| Cause of death | Death circumstance | `donor_DEATH_CIRCUM_DON` |
| Cause of death | Death mechanism | `donor_DEATH_MECH_DON` |
| Cause of death | Cause-of-death code | `donor_COD_CAD_DON` |
| Donation pathway | Legally brain dead | `donor_LEGALLY_BRAIN_DEAD` |
| Donation pathway | Donation after circulatory death progressed to brain death | `donor_DCD_PROGRESSED_BRAIN_DEATH` |
| Resuscitation | Cardiac-arrest downtime duration | `donor_CARDARREST_DOWNTM_DURATION` |
| Resuscitation | Cardiopulmonary resuscitation administered | `donor_CPR_ADMIN` |
| Resuscitation | Cardiopulmonary resuscitation duration | `donor_CPR_ADMIN_DURATION` |
| *Recipient features* | | |
| Demographic | Blood group | `recipient_ABO` |
| Demographic | Age at transplant | `recipient_AGE` |
| Anthropometric | Body mass index | `recipient_BMI_CALC` |
| Anthropometric | Body mass index at TCR | `recipient_BMI_TCR` |
| Substance-use history | Cigarette-use status | `recipient_CIG_USE` |
| Immunologic | Calculated panel-reactive antibody | `recipient_CPRA` |
| Immunologic | Peak calculated panel-reactive antibody | `recipient_CPRA_PEAK` |
| Renal function | Serum creatinine at TRR | `recipient_CREAT_TRR` |
| Comorbidity | Diabetes | `recipient_DIAB` |
| Diagnosis | Diagnosis code | `recipient_DIAG` |
| Renal support | Dialysis after listing | `recipient_DIAL_AFTER_LIST` |
| Respiratory support | ECMO at TRR | `recipient_ECMO_TRR` |
| Demographic | Ethnicity category | `recipient_ETHCAT` |
| Demographic | Ethnicity | `recipient_ETHNICITY` |
| Pulmonary function | $FEV_1$ at TRR | `recipient_FEV1_TRR` |
| Functional status | Functional status at TCR | `recipient_FUNC_STAT_TCR` |
| Functional status | Functional status at TRR | `recipient_FUNC_STAT_TRR` |
| Pulmonary function | FVC at TRR | `recipient_FVC_TRR` |
| Demographic | Sex | `recipient_GENDER` |
| Diagnosis | Diagnostic grouping | `recipient_GROUPING` |
| Hemodynamics | Cardiac output at TRR | `recipient_HEMO_CO_TRR` |
| Hemodynamics | Pulmonary artery diastolic pressure at TRR | `recipient_HEMO_PA_DIA_TRR` |
| Hemodynamics | Mean pulmonary artery pressure at TRR | `recipient_HEMO_PA_MN_TRR` |
| Hemodynamics | Pulmonary capillary wedge pressure at TRR | `recipient_HEMO_PCW_TRR` |
| Hemodynamics | Systemic blood pressure at TCR | `recipient_HEMO_SYS_TCR` |
| Anthropometric | Height, cm | `recipient_HGT_CM_CALC` |
| Anthropometric | Height, cm, at TCR | `recipient_HGT_CM_TCR` |
| Pulmonary vasodilator | Inhaled nitric oxide use | `recipient_INHALED_NO` |
| Initial waitlist data | Age at initial waitlist assessment | `recipient_INIT_AGE` |
| Initial waitlist data | Body mass index at initial waitlist assessment | `recipient_INIT_BMI_CALC` |
| Initial waitlist data | Height at initial waitlist assessment | `recipient_INIT_HGT_CM_CALC` |
| CAS component | Initial CAS biologic score | `recipient_INIT_MATCH_CAS_BIO_SCORE` |

*Table S1 continued*

| Domain | Feature | Input variable |
|---|---|---|
| CAS component | Initial CAS PTACC score | recipient_INIT_MATCH_CAS_PTACC_SCORE |
| CAS component | Initial CAS subtotal score | recipient_INIT_MATCH_CAS_SUBSCORE |
| CAS component | Initial CAS waitlist medical-urgency score | recipient_INIT_MATCH_CAS_WL_SCORE |
| Initial waitlist data | Initial candidate status | recipient_INIT_STAT |
| Initial waitlist data | Weight at initial waitlist assessment | recipient_INIT_WGT_KG_CALC |
| Hemodynamic support | Inotrope use at TCR | recipient_INOTROPES_TCR |
| Hemodynamic support | Inotrope use at TRR | recipient_INOTROPES_TRR |
| Life support | Life support at TCR | recipient_LIFE_SUP_TCR |
| Comorbidity | Malignancy at TCR | recipient_MALIG_TCR |
| Clinical condition | Medical condition at TRR | recipient_MED_COND_TRR |
| Transplant history | Number of previous transplants | recipient_NUM_PREV_TX |
| Respiratory support | Oxygen requirement | recipient_O2_REQ_CALC |
| Life support | Other life support at TCR | recipient_OTH_LIFE_SUP_TCR |
| Blood gas | Arterial $PaCO_2$ at TRR | recipient_PCO2_TRR |
| Pulmonary vasodilator | Prostaglandin E use at TCR | recipient_PGE_TCR |
| Surgical history | Prior cardiac surgery at TCR | recipient_PRIOR_CARD_SURG_TCR |
| Surgical history | Prior lung surgery at TRR | recipient_PRIOR_LUNG_SURG_TRR |
| Pulmonary vasodilator | Prostacyclin use at TCR | recipient_PROSTACYCLIN_TCR |
| Pulmonary vasodilator | Prostacyclin infusion at TCR | recipient_PROS_INFUS_TCR |
| Clinical status | Candidate status at TRR | recipient_STATUS_TRR |
| Diagnosis | Diagnosis at TCR | recipient_TCR_DGN |
| Diagnosis | Thoracic diagnosis | recipient_THORACIC_DGN |
| Respiratory support | Mechanical ventilator use at TRR | recipient_VENTILATOR_TRR |
| Respiratory support | Ventilatory support at TRR | recipient_VENT_SUPPORT_TRR |
| Anthropometric | Weight, kg | recipient_WGT_KG_CALC |
| Anthropometric | Weight, kg, at TCR | recipient_WGT_KG_TCR |

Abbreviations: ABO, blood group; BMI, body mass index; CAS, Composite Allocation Score; cPRA, calculated panel-reactive antibody; CPR, cardiopulmonary resuscitation; ECMO, extracorporeal membrane oxygenation; $FEV_1$, forced expiratory volume in 1 second; $FiO_2$, fraction of inspired oxygen; FVC, forced vital capacity; $PaCO_2$, arterial partial pressure of carbon dioxide; $PaO_2$, arterial partial pressure of oxygen; P/F, $PaO_2/FiO_2$; TCR, transplant candidate registration; TRR, transplant recipient registration.
